# Optimizing rabies vaccination of dogs in India

**DOI:** 10.1101/2023.04.10.23288318

**Authors:** Kim Cuddington, William H.B. McAuliffe

## Abstract

Dog vaccination is the key to controlling rabies in human populations. However, in countries like India, with large free-roaming dog populations, vaccination strategies that rely only on parenteral vaccines are unlikely to be either feasible or successful. Oral rabies vaccines could be used to reach these dogs. We use costs estimates for an Indian city and linear optimization to find the most cost-effective vaccination strategies. We show that an oral bait handout method for dogs that are never confined can reduce the per dog costs costs of vaccination, and increase vaccine coverage. This finding holds even when baits cost up to 10x the price of parenteral vaccines, if there is a large dog population or proportion of dogs that are never confined. We suggest that oral rabies vaccine baits will be part of the most cost-effective strategies to eradicate human deaths from dog-mediated rabies by 2030.

## Introduction

Rabies is a neglected tropical disease [1]that has the highest mortality rate of all known infectious agents [2]. Recent global initiatives aim at achieving zero human deaths from rabies by 2030 [3]. The majority of deaths occur in Africa and Asia, where free-roaming dogs are the primary means of transmission. India has a large free-roaming dog population, and probably accounts for 36% of human rabies deaths [4]. In this contribution, we use optimization techniques to show that the most cost-effective vaccination strategies for eradicating rabies in India will include vaccination of free-roaming dogs with oral rabies vaccine baits, which is not currently part of the management of rabies.

Rabies deaths are under-reported [5]. Incubation can take 1-3 months following infection, and the paralytic form of the disease is not widely recognized (e.g., [6]). In addition, although rabies is a notifiable disease in many countries where it is endemic, it has not been in all (e.g., the Indian government only recently announced that reporting would be required, [7]). Unfortunately, notifiability does not necessarily ensure effective surveillance either [8, 9]. The World Health Organization (WHO) suggests that an estimate of ~ 59,000 deaths per year, which attempts to account for under-reporting [10], is also likely an underestimate [11]. As a result, the estimate for India of 18,000 - 20,000 deaths per year [4], is probably too low.

The negative economic impact of rabies is relatively large. Hampson et al. [10]estimates there is a global economic burden of ~ USD $8.6 billion annually [1], with related welfare impacts, including deaths, at 3.7 million disability-adjusted human life years (DALYs) lost. About 29 million post-exposure prophylaxis treatments (PEP) are delivered each year [1], with 9 million in India [10], and this treatment comprises ~ 83% of the total rabies control budget in both Asia and Africa [12]. The disease is disproportionately found in poor rural populations [1], for whom treatment costs may be a heavy burden. Timely PEP is almost 100% effective in preventing death [13], but because of costs, currently estimated at USD $108 including travel and loss of income [1], some patients do not finish treatment. Livestock losses due to rabies are estimated at USD $62 million (CI $29 - $237 million, [10])and, again, may be disproportionately borne by those in poor rural areas.

Over 99% of cases of human rabies are caused by an infected dog bite [1]. The WHO suggests that dog vaccination is the most cost-effective strategy for preventing rabies in people, and reduces both human deaths and the need for PEP [1]. Programs in Tanzania, the Philippines, and South Africa [14]found that while costs per dog vaccinated varied (~$1.18 - $15.62 2012 USD), they were much lower than costs of PEP ($44.91 to $64.38 2012 USD). A recent cost comparison in Chad also suggests canine mass vaccination has approximately double the cost-effectiveness per DALY averted compared to PEP alone [15]. Investment in dog vaccination, however, accounts for less than 1.5% of the global economic burden of the disease, and until recently, large-scale dog vaccination activities in India accounted for less than 0.5% of the estimated economic burden from the disease [10].

The WHO, Food and Agriculture Organization (FAO), and the World Organization for Animal Health (OIE) have prioritized rabies as a model disease for a One Health approach. These agencies have launched the ‘United Against Rabies Forum’ that advocates and prioritizes investments in rabies control, and coordinates global efforts to achieve zero human deaths from dog-mediated rabies by 2030 [3]. However, these campaigns do not include financial pledges. Moreover, most charitable donations associated with rabies control are commitments to contribute to the costs of PEP in endemic regions (e.g., [5]), while investment in dog vaccination has been judged insufficient [16].

Annual vaccination of over 70% of the dog population can stop transmission and eventually lead to elimination if repeated over several years [17], while other strategies, such as culling, are less effective [18, 19]. Mass vaccination campaigns targeting dogs have been highly successful in many countries. For example, the U.S. was able to eliminate the canine rabies variant in the late 1970s and again in the 2000s. Widely used strategies for dog vaccination include central point vaccination and door-to-door vaccination. In central point vaccination (CP) dog owners bring their pets to a central location such as a veterinarian office. Door-to-door (DD) strategies are where teams move from home-to-home to vaccinate dogs that can be handled by their owners. These two strategies have only been successful at large scales where most dogs are responsibly owned (e.g., Latin America, [20]).

In countries like India, where there are many free-roaming dogs that may not be owned [21], catch-vaccinate-release techniques (CVR) have been employed. For animals that cannot be easily handled, CVR entails a team of people (4-7) capturing the animal in a net, injecting the vaccine, and then releasing. For example, in 2013, the charity Mission Rabies (https://missionrabies.com/) conducted synchronized mass dog vaccination campaigns in 12 Indian cities using CVR and vaccinated 54,227 dogs [22]. However, there is no example of a large-scale national campaign that relies primarily on CVR. Wallace et al. [23]and Gibson et al. [22]suggest that the labor force required for such a campaign is prohibitively large.

Instead, several authors promote oral rabies vaccine bait handouts (ORV) as a key strategy in the control of canine rabies where there are large populations of free-roaming dogs [24, 25, 23]. This method involves providing attractive oral rabies vaccine baits to animals that cannot be easily handled, observing consumption or removing the bait if rejected. The WHO has been recommending ORV as a complementary measure to reach inaccessible dogs since at least 1998 [26]. Moreover, oral baits dispersed in the environment have been used successfully in North America and Europe to control rabies in a variety of wildlife species, and have resulted in a net savings in disease control costs (e.g., [27]). India formally endorsed the use of ORV for a WHO-recommended dog oral vaccine, SAG2, in 2007, but the costs of commercial vaccine baits exceeded funding [25].

It is likely, however, that the total costs of ORV vaccination of free-roaming dogs will be lower than those of CVR. Mission Rabies conducted a pilot test of oral baiting in Goa, India in 2018, where they compared ORV and CVR using an empty bait construct [24]. The fixed cost of ORV was one quarter of CVR, and had a faster daily vaccination rate. ORV also increased the proportion of dogs accessible for vaccination across land use types, such as urban areas and rural villages. Further, staff reported that dogs were more likely to run away from CVR teams and alert nearby dogs by barking, while ORV teams reported that dogs were often attracted to the baits.

Gibson et al. [22]used a spreadsheet tool, originally created by Wallace et al. [28]to calculate the costs of canine vaccination campaigns which included ORV for Indian cities. We show how a simple optimization routine can instead use this same information to identify the best vaccination strategy for dog populations with different proportions of free-roaming animals, and for a range of possible bait costs. This technique can suggest cheaper strategies that may not have been considered by practitioners. For the scenarios examined, we find including ORV almost always improves cost-effectiveness.

## Methods

We used linear programming to determine the optimal combination of canine rabies vaccination methods that will minimize costs for a desired level of vaccination coverage, for a range of scenarios.

### Structure and size of dog population

Following [22, 28], we divided the total dog population into three categories: always confined (C), sometimes confined (SC), and never confined (NC). To determine if there were cost savings of ORV we compared optimal solutions where four different methods of vaccination were available (CP, DD, CVR, and ORV), or only the three standard methods (CP, DD, and CVR). The number of dogs and the proportion in each category in a region is most uncertain, so we varied the total population size and proportion of NC dogs to examine a range of scenarios. The usefulness of ORV will depend heavily on the number of NC dogs, and because of this, we merely divided the remaining population evenly between C and SC dogs.

### Vaccination costs

For each scenario, we used per dog vaccination costs as input to an optimization routine to identify strategies that minimized the final costs. Per dog vaccination costs were calculated from a selection of mean cost estimates proposed by [28]and specified for the city of Bangalore (see Supplementary material Bang Scen A - OBH - 11d.xlsx from [22]). In some cases, we introduced modifications (e.g., a doubling of the vehicle costs for CVR compared to DD/OVR which might be required with the larger team size). In other cases we estimated quantities that were not provided (see details in Appendix). Since there is considerable uncertainty regarding the exact price of oral baits for use in India [23], we allow the this cost to vary from $0.50 to $5.50 in our calculations.

We determined personnel costs by setting the team size required for each method as 1, 2, 2 and 4 people for CP, DD, ORV and CVR respectively. We then calculated the per dog costs using vaccination rates of 30 dogs/team/day for CP, DD, and CVR methods, and either the same rate for ORV (Table 1), or a rate 50 of dogs/team/day to reflect the faster handling rate of this method (a rate used in [22]). These rates are within the range of those measured in high density urban areas [22]. We then used these costs for the optimization procedure.

**Table 1:** Per dog vaccination costs calculated using estimates from [22] with differences noted in the Methods

| Item | Cost per dog (USD) |
| --- | --- |
| Vaccines (Parenteral) | 0.40 |
| Vaccines (Oral) | 0.50-5.50 |
| Syringes and needles (CP/DD/CVR) | 0.13 |
| Vaccination certificates (CP/DD) | 0.05 |
| Dog marking (CVR/ORV) | 0.03 |
| CP technicians (1 per dog) | 0.40 |
| CVR technicians (4 per dog) | 1.47 |
| DD/ORV technicians (2 per dog) | 0.87 |
| CVR driver | 0.27 |
| CVR vehicle rental & fuel | 2.00 |
| DD/ORV vehicle rental & fuel | 1.00 |
| CP/DD bite PEP (1 in 2,000) | 0.05 |
| CVR bite PEP (1 in 500) | 0.20 |
| ORV bite PEP (1 in 1,000) | 0.10 |

### Vaccination coverage

The vaccination coverage achieved by these methods is determined by both vaccine efficacy and accessibility of dogs to these various methods. Like Gibson et al. [22]we assume parenteral vaccination has the highest efficacy at 100% chance of rabies immunity, while ORV provides only an 80% chance of immunity. Vaccine accessibility is the probability that a given vaccination method can reach a dog of a given category (see Appendix for more discussion), and was taken from example values in the spreadsheets provided by [22](see Table 2). However, we also examined the effect of varying accessibility of NC dogs to ORV, and SC dogs to CP.

**Table 2:** Example of vaccination accessibility from [22]

|  | CP | DD | CVR | ORV |
| --- | --- | --- | --- | --- |
| C | 0.95 | 0.95 | 0.05 | 0.05 |
| SC | 0.80 | 0.80 | 0.70 | 0.95 |
| NC | 0.05 | 0.10 | 0.64 | 0.79 |

To get maximum possible vaccination coverage for a given method, we multiply the probability of being able to use a given vaccination method on each dog confinement category (Table 2) by vaccination efficacy. For example, ORV can reach 79% of NC dogs (Table 2), but since it has an estimated 80% efficacy once administered, this method has a maximum coverage of 63%. CVR has an accessibility of 64%, and parenteral vaccination provides a 100% immunity, so the maximum coverage rate remains quite similar at 64%.

### Optimization

To determine optimal vaccine delivery solutions, we use linear programming. This approach has been used for other similar health care problems (e.g., [29, 30]). Our task is an example of a linear transport problem with multiple constraints.

The objective function of a linear programming problem describes the main objective of the decision-maker. In this case, we wish to minimize the per dog costs of vaccination, while maintaining a minimum level of vaccination coverage (see below *Constraints to achieve rabies vaccination targets*). To do this, we need to find the optimal number of dogs to vaccinate using each vaccination method.

Our objective function is:

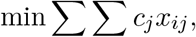

where *c*_*j*_ is the cost for each vaccination method, as described above, and *x*_*ij*_ is the number of dogs vaccinated in each of the three categories, *i* by one of the four methods *j*. Optimal solutions were found using lpSolve, an R [31]interface to the freely available software lp_solve (version 5.5, https://lpsolve.sourceforge.net/5.5/).

### Constraints to achieve rabies vaccination targets

In addition to minimizing costs, we added constraints to ensure the selected solution met a minimum vaccination target for disease transmission to be halted or at least slowed. For C and SC dog categories, we use 70% as the minimum annual vaccination coverage required for rabies control [17]. We add this constraint as: L. *x*_*ij*_*v*_*ij*_ ≥ 0.7*d*_*i*_, where *x*_*ij*_ is the number of dogs in category *i* vaccinated by method *j, d*_*i*_ is the number of dogs in each category in the population, and *v*_*ij*_ is the maximum coverage of method *j* on dog category *i*, where this quantity includes both accessibility and vaccine efficacy (see above *Vaccination coverage*).

Given the example vaccine efficacy and accessibility of ORV and CVR provided by [22]for Bangalore, this 70% vaccination target cannot be met for NC dogs (although this coverage has been met in some studies [22]). However, rabies may have relatively low transmission rates, such that in some populations lower vaccination coverage may be sufficient to substantially reduce economic and DALY impacts. For example, Fitzpatrick et al. [32]predict an 88% reduction in annual human rabies deaths for an ongoing program of canine vaccination that reaches ~ 13% of the overall dog population. We therefore set the vaccination target for NC dogs at 60%, to fall slightly below the maximum possible coverage for CVR and ORV methods for the example values in [22].

### Final costs

We generated optimal solutions (see example in Table 3) for each combination of oral bait price (ranging from $0.5 to $5.50), proportion of NC dogs (ranging from 0.05 to 0.99), over a range of total dog population sizes (5,000 - 150,000), with target vaccination thresholds of 70% for C and SC dogs, and 60% for NC dogs, where this lower value for NC dogs is close to the maximum possible for either ORV or CVR.

**Table 3:** Optimal vaccination strategy for a population of 50,000 dogs with 48% NC, oral bait cost of $2.50 with other costs as given in Table 1 and vaccination accessibility as given in Table 2.

|  | C | SC | NC |
| --- | --- | --- | --- |
| Total dogs | 13000 | 13000 | 24000 |
| Target coverage | 70% | 70% | 60% |
| **Number of vaccinations needed** |  |  |  |
| CP | 9579 | 11375 | 0 |
| DD | 0 | 0 | 0 |
| CVR | 0 | 0 | 0 |
| ORV | 0 | 0 | 22785 |

After an optimal vaccination strategy was identified, we then calculated the cost of a 30 day campaign to yield a final per dog cost. Since we assumed fixed costs did not vary significantly for different optimal strategies that used only CP, DD and CVR, these were not included. However, we include one additional fixed cost for campaigns that included ORV: $10,000 for an information campaign specific to oral baits (see Appendix). As a result, a strategy identified by the optimization as having the lowest per dog vaccination cost could have a higher final per dog cost than other strategies.

Where solutions were possible (see above regarding *Constraints to achieve rabies vaccination targets*), we then compared the optimal solution obtained when the four different vaccination methods were available to that where only the three standard methods were used, in order to determine if costs were lowered by incorporating ORV.

## Results

In general, the final cost per dog was reduced with use of ORV if there was a large number of NC dogs. For example, linear optimization suggests that 25,000 NC dogs in a total population of 50,000, ORV use will yield lower per dog costs until a fairly high oral bait cost of $3.85 (Figure 1a). However, lower CVR vehicle costs will push this price threshold lower (Figure 1b).

**Figure 1:**
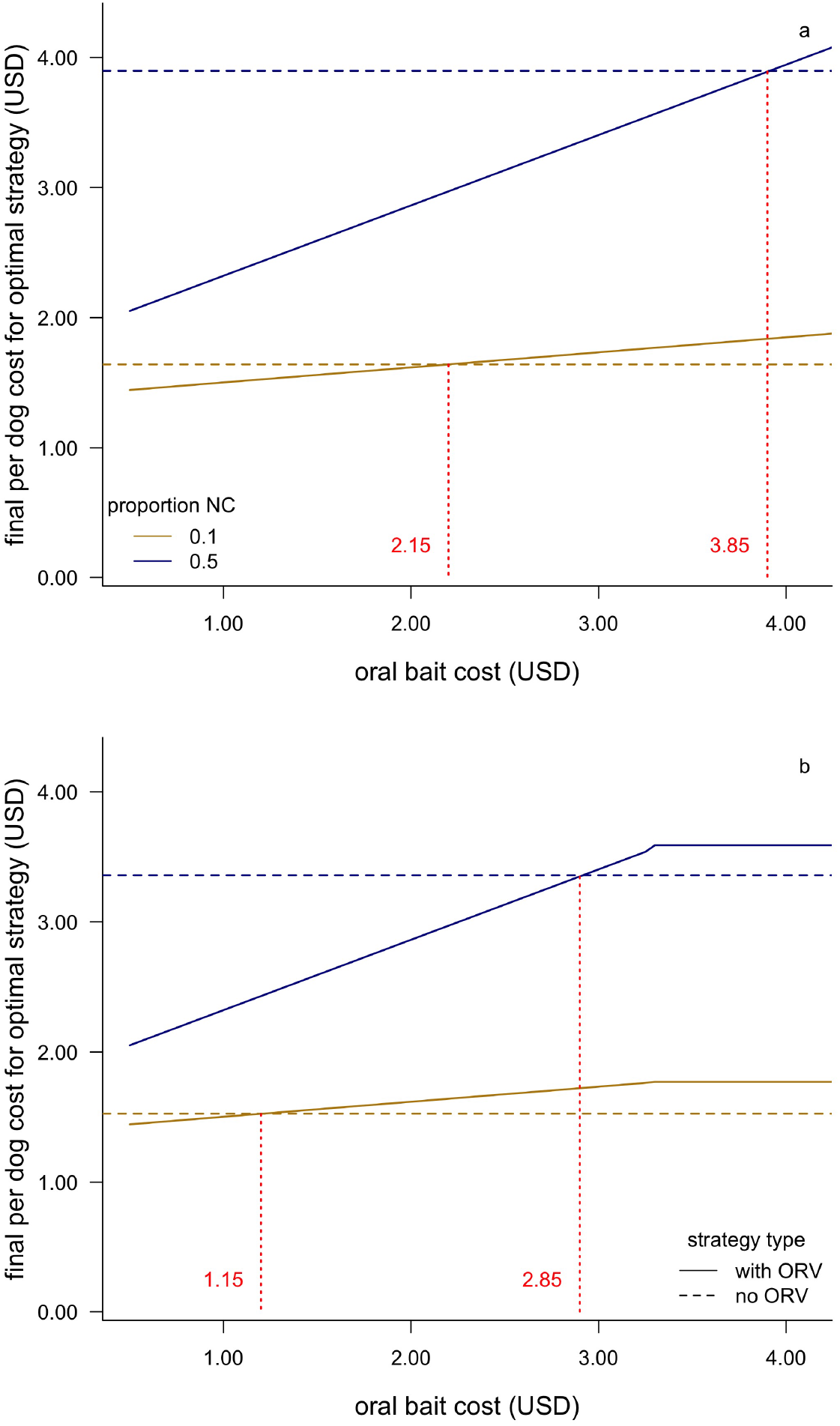
Final per dog costs for vaccination campaigns with and without the use of oral rabies vaccine bait handout (ORV) for a dog population of 50,000, where the number of never confined dogs (NC) is either 50% or 10% of the population, and the catch-vaccinate-release (CVR) vehicle costs are either as given in Table 1 (a), or one-half this value (b). Horizontal dashed lines give the final per dog cost without ORV, while solid lines show how final per dog costs increase with bait cost. The intersection of the lines gives the bait cost at which the strategies have the same final per dog costs

We find a similar price threshold across a range of proportion NC dogs in this fixed population size (Figure 2). Unless the oral bait price is greater than roughly 10x the price of the parenteral vaccine, or the proportion of NC dogs is less than ~0.2, ORV use gives lower costs. With a larger population size, or a higher proportion of NC dogs, this price threshold is even higher (e.g., $4.15 for 150,000 dogs, see Figure 3). However, for very small proportions of NC dogs (e.g., <0.1) there may be no cost advantage unless the total population is quite large.

**Figure 2:**
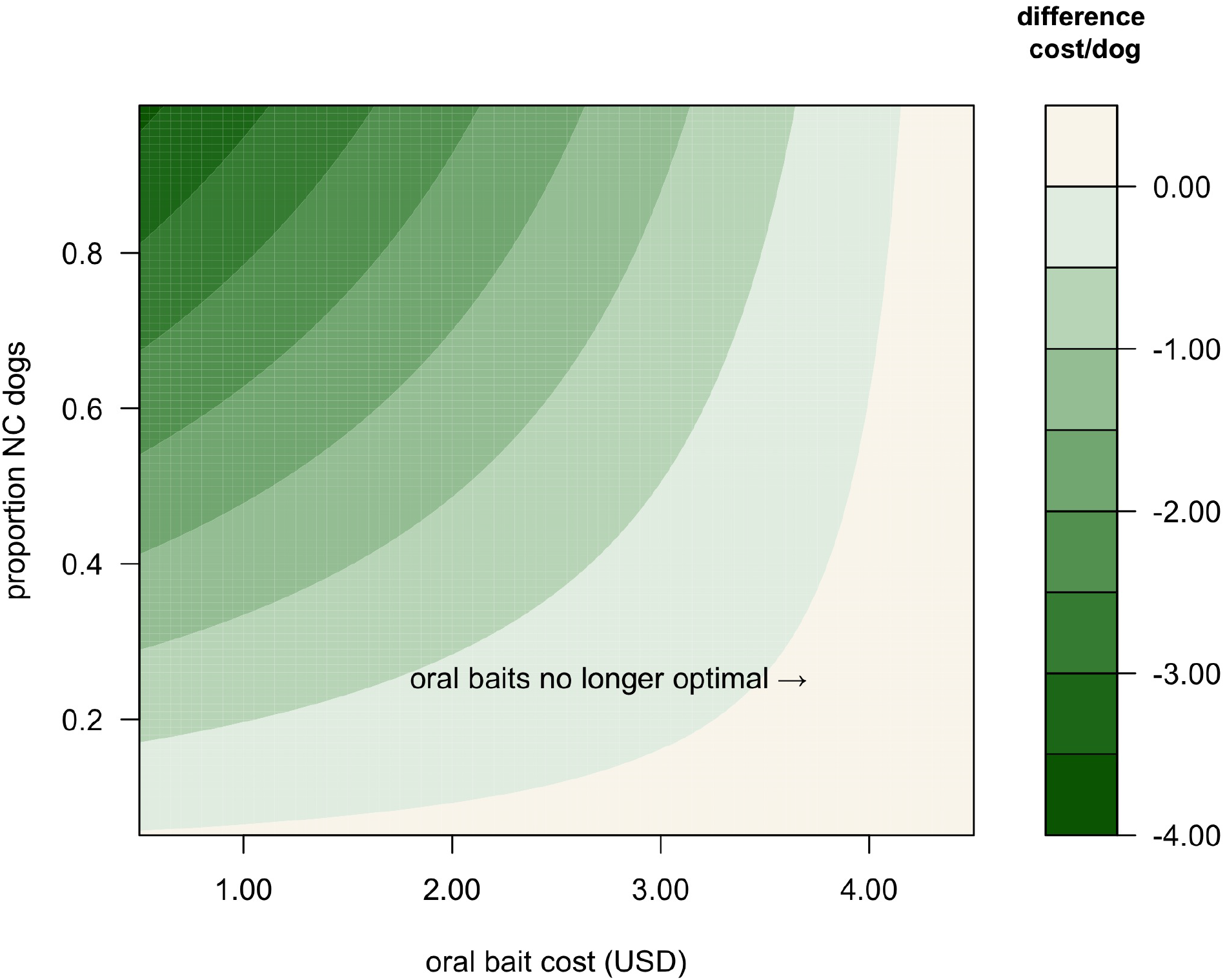
Difference between final per dog costs for optimal vaccination strategies without and with oral rabies vaccine handout (ORV) for a total dog population of 50,000 with varying proportions of never confined (NC) dogs and oral bait cost, and other costs as given in Table 1. Negative values indicate the reduction in final per dog cost when using OVR

**Figure 3:**
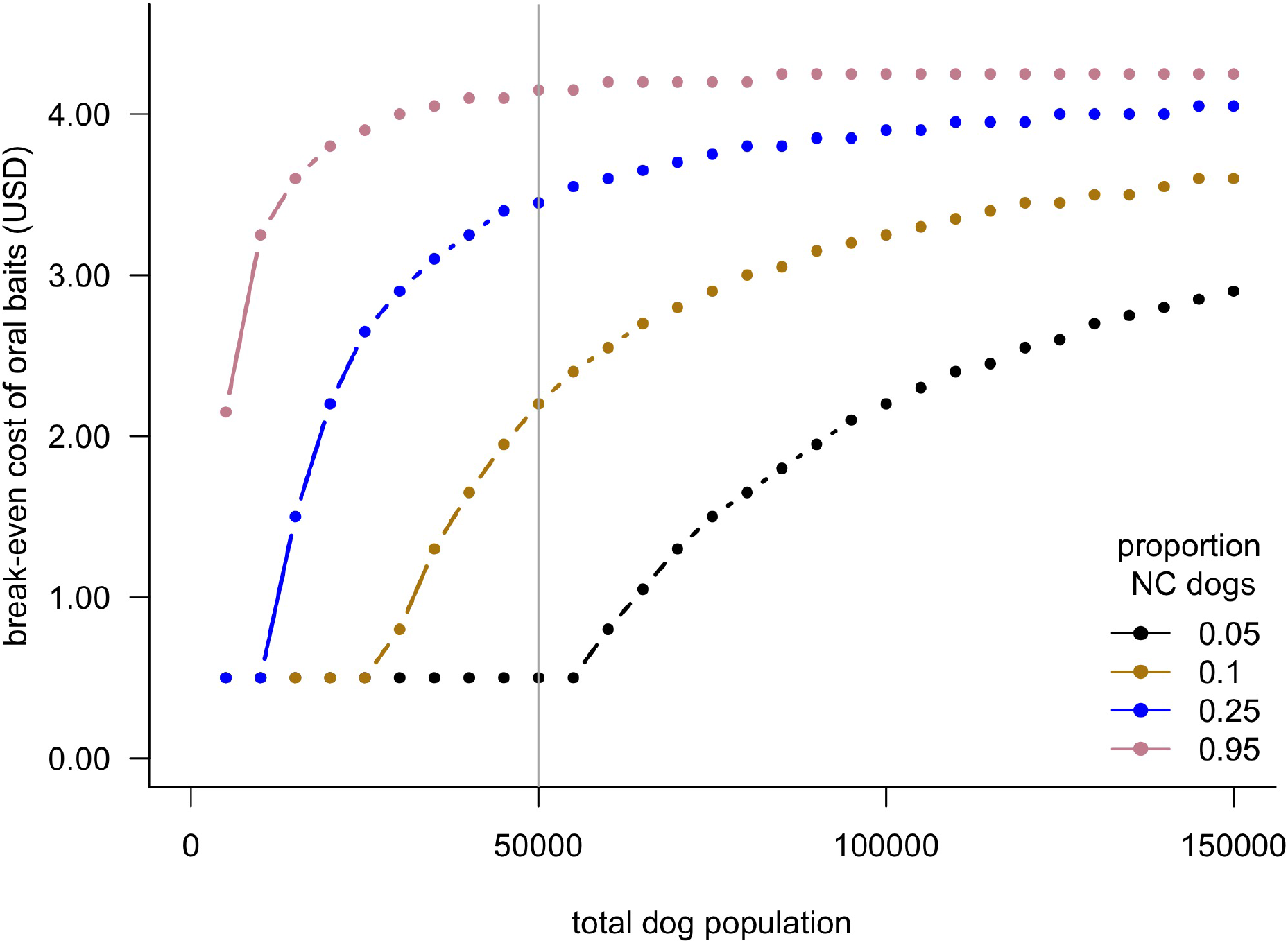
Maximum cost per oral bait at which there would no longer be a cost advantage of using oral rabies vaccine bait handout (ORV) for costs as given in Table 1, accessibility as in Table 2, and a range of total dog population sizes and different proportions of never confined (NC) dogs. Vertical line shows outcomes for a population size of 50,000 dogs used for other figures

We categorized the optimal solutions across the range of oral bait price and proportion of NC dogs for a intermediate fixed population size of 50,000 dogs. There are two major categories of solution for NC dogs, but CP vaccination is always suggested for C and SC dogs for the vaccination accessibilities in Table 2. If the oral bait cost is less than the price threshold the optimal strategy is to always use ORV for NC dogs, and CVR otherwise.

Changes to ORV accessibility will change the optimal solution. Unsurprisingly, optimal solutions are more likely to include ORV for higher accessibility and lower cost (Figure 4).

**Figure 4:**
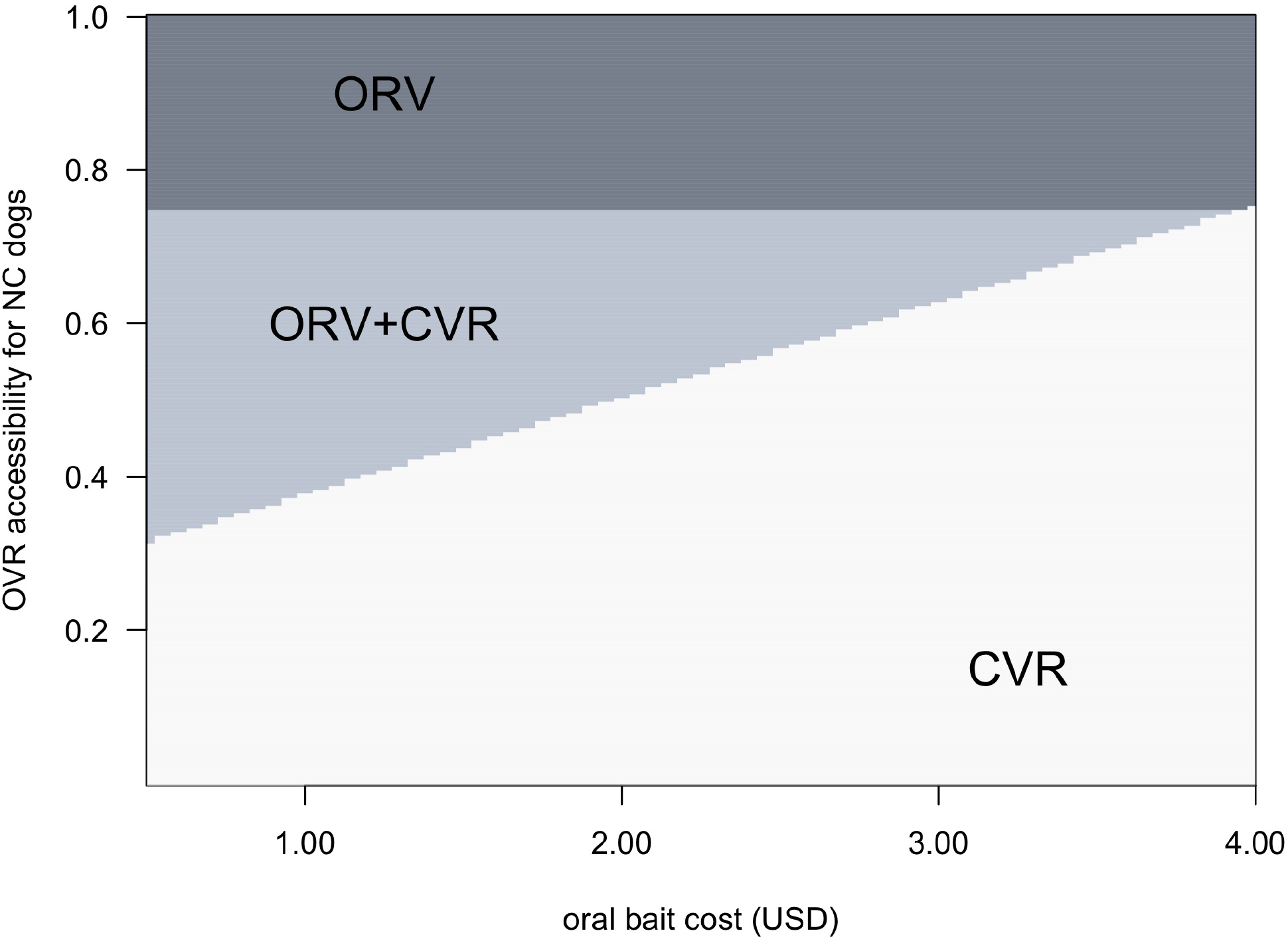
Most cost-effective vaccination strategies for never confined (NC) dogs with the constraint of 60% vaccination coverage. Shaded areas and text indicate whether the optimal strategy is to use oral rabies vaccine bait handout (ORV), catch-vaccinate-release (CVR) or a mixed strategy as oral bait cost and OVR vaccination accessibility vary. Other costs as given in Table 1, and CVR accessibility is fixed at 0.64. We show the scenario where NC dogs comprise 50% of a total dog population of 50,000

Interestingly, ORV is part of the optimal solution even when accessibility is low (e.g., 0.5) for moderate bait cost ($2.50). For example, as ORV accessibility ranges from 0.0 - 0.99 for a population of 50,000 dogs, we find three different optimal strategies: ORV alone when accessibility is greater than 0.75, a combination of ORV and CVR when accessibility is lower and oral bait cost is less than $3.85, or CVR alone for lower accessibility and higher costs.

Similarly, the optimal strategy for other dog categories can vary with changes to vaccination method accessibility. When we allow the CP compliance rate for owners of SC dogs to vary, DD is preferred for low accessibility (Figure 5). We find there are even solutions where use of low cost ORV may be optimal for this category, when the daily vaccination rate for this method is higher than DD.

**Figure 5:**
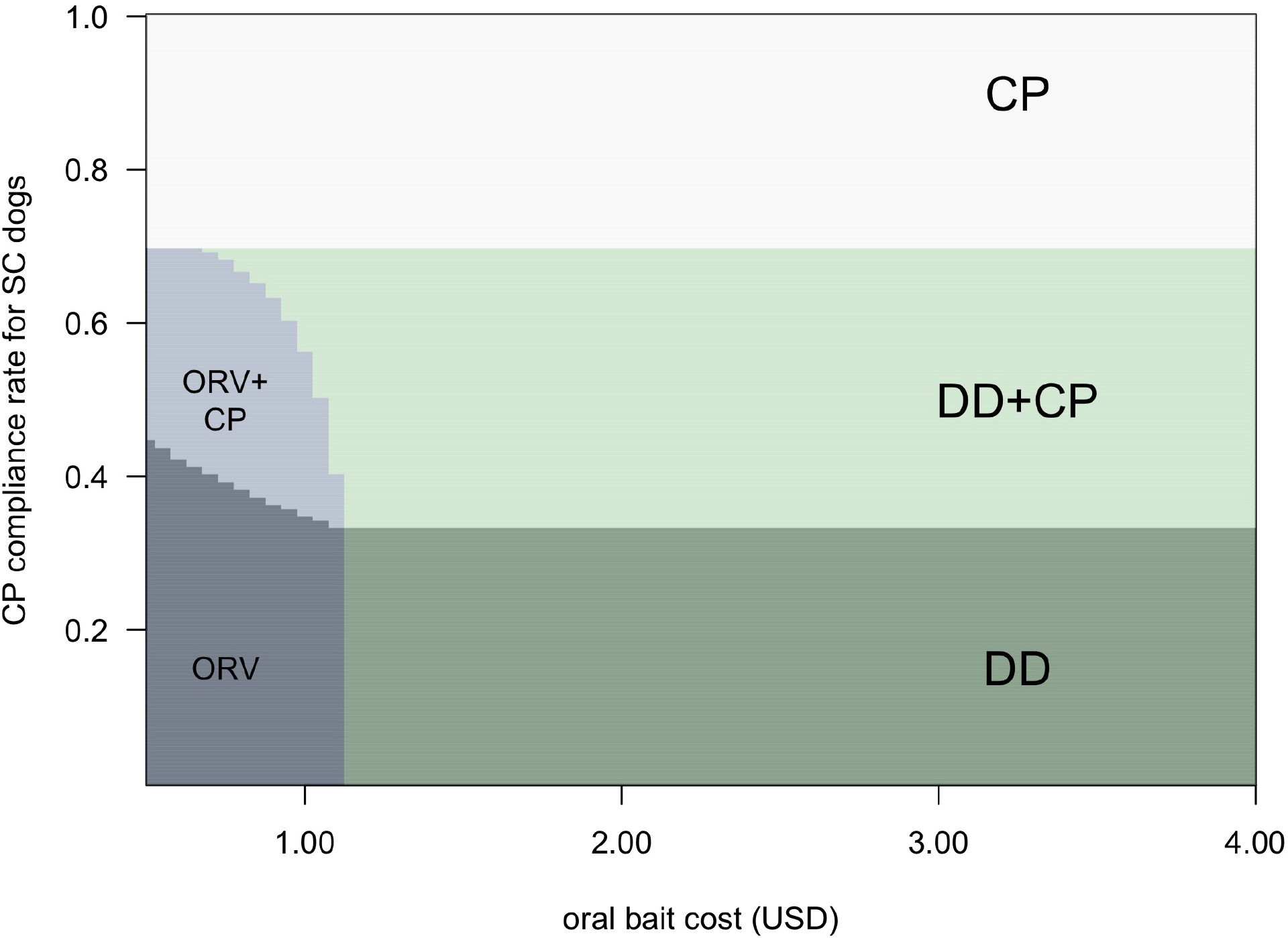
Most cost-effective vaccination strategies for sometimes confined (SC) dogs with the constraint of 70% coverage. Shaded areas and text indicate whether the optimal strategy is to use central point (CP), door-to-door (DD), oral rabies vaccine bait handout (ORV) or mixed strategies as oral bait cost and CP vaccination accessibility vary, where the ORV vaccination rate is faster than DD and CP at 50 dogs/team/day. We show the scenario where SC dogs comprise 50% of a total dog population of 50,000

## Discussion

A renewed commitment to achieve zero human deaths from dog-mediated rabies in India [7]requires effective vaccination solutions. However, this country faces the difficult problem of vaccinating large populations of free-roaming dogs. In cities like Bangalore, recent estimates suggest that the free-roaming population may be as large as 300,000 dogs (Worldwide Veterinary Service Centre, as reported by [33]). Our analysis indicates that for dog populations like these, oral rabies vaccine bait handout (ORV) may minimize costs, while still meeting reasonable vaccination coverage targets. Importantly, we find that ORV can offer significant cost-savings even when the baits themselves cost significantly more than parenteral vaccines, and have lower efficacy.

Using cost estimates from previous work [28, 22], we show that ORV becomes more cost-effective as the number of never confined (NC) dogs increases because the only other feasible method for these dogs, catch-vaccinate and release (CVR), usually has higher personnel and equipment costs for the same vaccination coverage. We examined a wide range of per unit prices for oral baits, assuming that initially baits would be imported at high cost, but may have lower costs with future domestic manufacture. In 2020, Wallace et al. [23]suggested a price range of $2.00-$4.00 USD while Gibson et al. [22]examined a range from $1.50-$2.50. We find that for some scenarios, even prices almost as high as 10x that of parenteral vaccines (~$3.85) can offer a cost-savings.

We note that all cost estimates included here are examples only and will likely vary widely from location to location. We expect the general trend of solutions to hold as long as the ratio of various costs remains similar. For example, reducing the vehicle costs of CVR by 50% still suggests that there is a bait price threshold below which ORV will be the best solution. However, if a particular cost category changes significantly relative to others, optimal strategies may change such that ORV is no longer part of the solution.

When it is suspected that the number NC dogs is small (e.g., <10% of population of 50,000 dogs), it is less clear if ORV should be employed. Unfortunately, dog population estimates are usually poor. There is often little survey data, and estimates are frequently created by using a fixed proportion of the human population. Photo mark-recapture data collection seems quite promising (e.g., [34])and could be used before the design of a vaccination campaign to estimate the size of free-roaming populations more accurately. Such methods may reveal that different areas in the same urban community have quite different population structures [35], so that more effective methods can be targeted for specific areas, again using optimization (e.g., [30]). However, for large numbers of NC dogs it seems clear that ORV will usually be the best option.

Optimal solutions were determined in part by the accessibility of dogs to different methods because of the requirement to meet vaccination coverage targets. Accessibility estimates will also vary widely with location. While we found solutions included an increased use of CVR as ORV accessibility decreased, we strongly suspect that low accessibility to ORV methods is correlated with low accessibility to CVR methods, except perhaps in the case of low bait palatability. We also note that it will be quite difficult to meet 70% vaccination targets for NC dogs with either of these methods, although the recent successes in Goa are inspiring [36]. Therefore, we suggest the efficacy of lower targets for reducing the human burden of disease should be further investigated.

Other vaccination accessibility changes can lead to optimal strategies that use ORV for different dog categories. In rural India, Tiwari [21]suggests that dogs are mostly “partially” owned, meaning that a household may claim ownership but not consider themselves responsible for the animal’s vaccination and veterinary care. In Bangalore specifically, there is a higher density of free-roaming dogs in areas with a higher human density and lower average income. Households in this area are also more likely to feed free-roaming dogs [35]. This scenario both increases risks of rabies transmission, and potentially makes it less likely household-associated dogs will be transported to a central location for vaccination, or even encountered during door-to-door vaccination efforts. Owner-driven vaccination programs used elsewhere (e.g., [20])may not be as effective in these regions. For low CP compliance rates, or low DD probability, optimal solutions can employ low cost ORV for these dogs.

Finally, methods not considered here may offer better cost savings. For example, mobile CP methods, where vaccination centers on vehicles move through neighborhoods, combined with DD, may be a better option when ORV costs are high. For fractious dogs, or staff that have not been trained in injection, oral bait handout with a door-to-door access method is another option that may be a reasonable choice. It may even be possible to combine oral contraceptives with the oral rabies vaccine bait, to simultaneously reduce population turnover.

In conclusion, pilot projects and analyses have previously suggested that oral baits will make a valuable contribution to India’s campaign to eliminate rabies [24, 22]. We use optimization to demonstrate that even if oral baits are considerably more expensive and less effective than parenteral vaccinates, they may still reduce costs in most scenarios involving free-roaming dogs, while providing very good vaccination coverage. We expect that charitable efforts aimed at providing low cost ORV may be more cost-effective in reducing the human burden of disease than additional efforts directed at PEP.

## Data Availability

This is a modelling study which has generated no data. Code is available at https://github.com/kcudding/rabies.

## Supplementary material

See Appendix.

## Data availability statements

All code used to generate our results can be found on github (see https://github.com/kcudding/rabies). In addition, the code requires spreadsheets in the supplementary material from [22] (http://www.mdpi.com/2414-6366/5/1/47/s1.)

## Appendix 1: Detailed methods for Cuddington and McAuliffe, Optimizing rabies vaccination of dogs in India

### Model formulation

We use linear optimization on per dog vaccination costs to identify cost-minimizing strategies with the constraint that the solution maintains the desired vaccination coverage, for a range of scenarios (see main text).

### Detailed vaccination costs

Some costs for consumables in Gibson et al. [1]were already provided on a per dog cost (e.g., vaccines, syringes, certificates and dog marking). Costs that were not be expressed per dog included vehicle rentals, equipment and staff wages that would scale with a given number of vaccinations (e.g., the costs of CVR include among other things, the cost of renting a van, the budgeted expense of treating employees that received a bite, and the salary of a driver).

Using the estimated daily wage, team size and vaccination rates (see main text) we can then calculate the per dog cost of personnel. Naturally these costs may vary wildly with location, but we confirmed that the rates used by Gibson et al. [1](i.e. $12, $13, $11, $13 daily wage for technicians using CP, DD, CVR and OVR vaccination respectively, and $8 for the driver) were reasonable (about double the current minimum wage for skilled worked in the state of Karnataka, [2]).

The per dog rate of other expenses such as CP/DD Bite PEP can be calculated from the rates provided (1 in 2,000). A separate rate for ORV was not provided, but we assumed the same cost as CP/DD PEP with a higher rate (1 in 1,000), but a lower rate than CVR PEP (1 in 500). We assumed that vaccination certificates would only be provided for CP and DD dogs, and dog marking (e.g., paint or similar) would be completed only for CVR and ORV dogs (see Table 1).

While DD and ORV vaccination requires a team of two and minimal equipment than can be transported on a small vehicle like a moped, CVR requires a lot of equipment and a vehicle that can transport 4-5 people. Therefore unlike Gibson et al. [1], we differentiate the vehicle rental costs and assume that the larger vehicle for CVR will cost 2 times more ($30 USD/day) than a vehicle required for DD and OBR ($15/day), and will require twice the amount of gasoline. We believe this is an underestimate of required vehicle costs, since our informal online search of rental agencies suggests moped rentals will range from $3 to $7 USD per day, passenger cars $25-85 and larger minivan ~$100. Therefore, while not included in this analysis, if mopeds could indeed be used for DD and ORV, and a larger vehicle such as a minivan was needed for CVR, the costs would differ by a factor of 10. Note that when the faster vaccination rate of 50 dogs/team/day for ORV is used, the per dog cost of personnel and vehicle rental for DD and ORV methods will differ from Table 1.

Wallace et al. [3]notes that the WHO suggests the use of oral baits requires information campaigns, as well as surveillance systems capable of detecting unintended vaccine exposures, which could add additional unknown costs to such a campaign. This recommendation for surveillance seems to relate more to the case where oral baits are broadcast in the environment rather than handed out to individual dogs. In addition, recent developments in these vaccines make adverse effects on accidental exposure very unlikely [4]. Nonetheless, we included one additional fixed cost for vaccination campaigns that included oral baits: $10000 for dissemination regarding the deployment of oral baits. This value was based on estimates from Gibson et al. [1]for a more general advertising campaign, and included costs for 10000 pieces of printed information and 30 days of radio or car loudspeaker announcements.

There are no differences in costs noted by these authors with respect to using the different methods for different classes of dogs. Using these estimates we obtain a per dog vaccination cost of $1.03, $2.50, $5.36-$6.36, for CP, DD and CVR methods respectively, and a range of $1.75-$7.50 for ORV. Thus, the cost of central point vaccination (CP) for a never-confined dog (NC), is given as the same for an always-confined (C) dog. The difference in using these methods for different categories is accounted for in an accessibility metric (see below *Detailed vaccination method coverage*).

### Detailed vaccination method coverage

We use the vaccine accessibility values provided by Gibson et al. [1]for the city of Bangalore (Table 2), but 1. the vaccine accessiblity values will depend on the particular location, and 2. their meaning lies in the exact method of implementation. For example, Gibson et al. [1]indicates that CP and DD campaigns will have a higher probability of reaching C and SC dogs, and estimates a 5% chance of vaccinating an always confined (C) dog with an oral bait handout. However, it is of course entirely possible to have a door-to-door campaign that employs oral baits instead of injections, and it may be cheaper and more effective to do so if trained personnel to administer injections are too costly, or the probability of owners responding to calls to attend CP vaccination locations is too low. We allow NC ORV and SC CP vaccine accessibility to vary in our analysis to address the first point, but have not included more hybrid strategies such as the use of oral baits in a door-to-door campaign.

In the main text we note that it is not possible to meet a 70% coverage target with the values provided by [1], and so set the coverage target for NC dogs at 60%. We note that another option is to set a 70% vaccination target for the combined transmission category of free-roaming dogs which includes both NC and SC dogs, as well as 70% of C dogs. Solutions that meet this constraint can only be found where NC dogs comprise <50% of the population, if only one vaccination attempt is made per dog, and the remaining dog population is divided evenly between SC and C dogs.

### Optimization with probabilistic constraints

We also investigated solutions where the probability of reaching the specified vaccination constraints was met with either a 20% probability or a 70% probability, on the grounds that lower probabilities of meeting vaccination targets may be sufficient for significant positive impact as suggested by Fitzpatrick et al. [5].

To incorporate a probabilistic approach to meeting vaccination targets, we use a chance constraint optimization procedure [e.g., 6]. We impose the constraint that for each dog category, *i*,

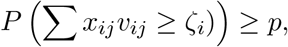

where *ζ*_*i*_ is the number of vaccinated dogs required to achieve the desired vaccination coverage for that category (e.g., 0.7*d*_*i*_), and p is a given probability.

Then, we can relax the probabilistic problem into the equivalent deterministic problem by using the appropriate probability density function and substituting the left hand side of the constraint with a deterministic expression. If we assume that *ζ*_*i*_ is normally distributed with mean *µ*_*i*_ and variance 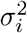, we can transform the constraint for each i into

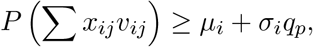

where *q*_*p*_ is the p-quantile of the standard normal distribution. We then assume that the dog population estimates were normally distributed with means as previously indicated and variance set as 20% of the mean. After this, we then solve as usual for a given p. However, we note that issues with convexity and stability can mean that small changes in the actual density function could cause major changes in the optimal solution.

However, introducing probabilistic constraints on meeting the vaccination targets did not introduce any differences in the optimal strategies. As expected, achieving a higher probability of success requires vaccinating more dogs. For instance, in a population of 50,000, we will need to vaccinate ~10% more dogs (5500-6400) to have 70% probability of meeting our coverage targets, versus having only a 20% probability of doing so.

